# Alzheimer’s disease blood-based biomarkers in the Boston Latino Aging Study (BLAST): Associations with cognition and depressive symptoms

**DOI:** 10.64898/2026.10.13.26365304

**Authors:** Catarina Tristão-Pereira, George Silva, Kelly A. Lopez, Vivian Ku, Amanda Gonzalez, Jairo E. Martinez, Averi Giudicessi, Meredith Rowe, Sandry Garcia, Marta Gonzalez Catalan, Daniel G. Saldana, Jennifer Gatchel, Liliana A. Ramirez-Gomez, Yakeel T. Quiroz

## Abstract

**Background:** Blood-based biomarkers (BBMs) may facilitate early detection of Alzheimer’s disease (AD), yet data in Latino populations remain limited. We examined associations of plasma biomarkers with cognition and depressive symptoms in Spanish-speaking Latino older adults enrolled in the Boston Latino Aging Study (BLAST).

**Methods:** Baseline data from 122 adults aged ≥55 years without dementia were analyzed. Plasma Aβ42/40, brain-derived phosphorylated tau (p-tau) 217, neurofilament light chain (NfL) and glial fibrillary acidic protein (GFAP) were measured using NULISA. Cognition was assessed with the Latin American Spanish version of the Preclinical Alzheimer’s Cognitive Composite (LAS-PACC5) and depressive symptoms with the Geriatric Depression Scale (GDS). Regression models were adjusted for age, sex, and education.

**Results:** Cognitively impaired participants had higher p-tau217 and GFAP than cognitively normal participants. Higher p-tau217 and GFAP were associated with poorer LAS-PACC5 performance, while higher NfL was associated with greater depressive symptoms. Aβ42/40 showed no significant associations.

**Conclusions:** Distinct biomarker associations with cognitive and emotional functioning support further longitudinal evaluation of BBMs for risk characterization in Latino older adults and potential disease monitoring applications.

## Introduction

Alzheimer’s disease (AD), the leading cause of dementia, affects approximately 7.4 million older adults in the United States. With the emergence of disease-modifying therapies, there is a growing need for accessible and scalable approaches to identify AD pathology early in the disease course[1]. Blood-based biomarkers (BBMs) offer a minimally invasive approach to detecting biological processes associated with AD [2]. Among these, phosphorylated tau (p-tau), particularly p-tau217, is strongly associated with cerebral amyloid and tau pathology, while the plasma amyloid-beta 42/40 (Aβ42/40) ratio reflects cerebral amyloid burden, although their performance may vary across assays and populations[3]. Neurofilament light chain (NfL) and glial fibrillary acidic protein (GFAP) provide complementary information about neuroaxonal injury and astrocytic activation, respectively, although neither is considered specific to AD [2]. Because changes in some BBMs can be detected during preclinical and early symptomatic stages and are associated with subsequent cognitive decline[4], they have considerable potential for early detection, risk stratification, and disease monitoring.

Despite these advances, most BBM research has been conducted in predominantly non-Latino White cohorts, limiting understanding of their performance and clinical relevance in populations historically underrepresented in AD research [5]. This limitation is particularly important for Latino older adults, a rapidly growing segment of the U.S. aging population that is expected to experience a substantial increase in AD and related dementias (ADRD) and yet remain underrepresented in biomarker studies and clinical trials [6,7]. Emerging research nevertheless supports the relevance of BBMs in this population. Among Hispanic/Latino adults, higher p-tau181/p-tau217 and NfL have been associated with poorer cognitive performance and functional impairment[8]. Furthermore, longitudinal studies of Mexican American older adults have linked higher GFAP and NfL to cognitive decline and identified GFAP as a predictor of incident dementia[9,10]. Together, these findings support the potential value of BBMs in Latino populations while emphasizing the need for further validation across diverse Latino samples.

Emotional functioning may also be important when evaluating biomarkers of cognitive aging. Latino older adults have been reported to experience high levels of depressive symptomatology [11], and depressive symptoms represent an important correlate of cognitive decline in older age. Examining cognition and depressive symptoms together may therefore provide a broader understanding of how AD-related and neurodegenerative biomarkers relate to cognitive and emotional functioning among Latino older adults.

To address these gaps, we established the Boston Latino Aging Study (BLAST), an ongoing longitudinal observational cohort of Spanish- and Portuguese-speaking Latino adults aged 55 years and older. BLAST was designed to characterize cognitive and emotional functioning and their biological correlates among Latino older adults without dementia. The present study reports initial baseline findings from Spanish-speaking BLAST participants and examines cross-sectional associations of plasma Aβ42/40, brain-derived p-tau217, NfL, and GFAP with cognitive performance and depressive symptoms. We hypothesized that biomarker profiles indicative of greater AD-related pathology or neurodegeneration would be associated with poorer cognitive performance and greater depressive symptoms. These baseline analyses provide an important foundation for evaluating the utility of BBMs in Spanish-speaking Latino older adults and for future longitudinal investigations within the expanding BLAST cohort.

## Methods

### Participants and general procedures

BLAST includes adults aged 55 years or older who are monolingual or bilingual speakers of Spanish, Portuguese, and/or English. Exclusion criteria include a diagnosis of dementia, a history of major neurological or psychiatric disorders (e.g., bipolar disorder or schizophrenia), or other conditions that could significantly affect cognition. The present analyses were restricted to Spanish-speaking participants who had completed a baseline study visit at the time of data extraction. In this study, we included participants who provided sociodemographic information, including age, sex, and educational attainment, underwent blood collection, and completed self-reported medical questionnaires and cognitive testing at baseline. Participants provided both written and verbal consent for participation, and all study-related procedures were approved by the Mass General Brigham Human Research Committee.

### Blood sample collection and processing

Fasting blood samples were collected from each participant at the MGH Clinical and Translational Research Unit (CTRU) in Boston, Massachusetts. Plasma was separated, divided into 1-mL aliquots, and stored at −80°C until biomarker analysis. Plasma analytes were measured using the NULISA platform and included Aβ40, Aβ42, p-tau217, NfL, and GFAP. These biomarkers were selected to capture complementary biological processes relevant to AD and related neurodegenerative processes, including Aβ and tau pathology, neuroaxonal injury, and astrocytic activation. NULISA Protein Quantification (NPQ) values[12], obtained after normalization to the assay’s internal controls and log2 transformation, were used for analysis. The Aβ42/Aβ40 ratio was additionally calculated as a measure more closely associated with cerebral Aβ pathology. Values outside Q1-4xIQR and Q3+4xIQR were considered outliers and excluded.

### Clinical and Cognitive measures

All neuropsychological assessments were administered in Spanish by trained bilingual psychometricians. Cognitive status was defined using as cutoff the average Mini-Mental State Examination (MMSE) score among individuals with Clinical Dementia Rating (CDR) of 0 (MMSE =23.3). Global cognitive functioning was assessed using the Latin American Spanish version of the Preclinical Alzheimer’s Cognitive Composite (LAS-PACC5) [13], a composite z-score derived from performance on the MMSE, the NEUROPSI Atención y Memoria 3rd edition Logical Memory Delayed Recall and Category Fluency (Animals) subtests, Oral Symbol Digit Modalities Test (SDMT), and the Free and Cued Selective Reminding Test (FCSRT). Depressive symptoms were assessed using the 30-item Geriatric Depression Scale (GDS)[14].

### Statistical analyses

We conducted descriptive statistics for all variables. We used analysis of covariance (ANCOVA) adjusted for age, sex and years of education to estimate group differences in plasma biomarkers between cognitively unimpaired and cognitively impaired participants. We then conducted linear regressions to examine the association of plasma biomarkers with LAS-PACC5 and GDS scores across the whole sample, adjusting for age, sex and years of education. To facilitate comparison across biomarkers, we standardized plasma levels using z-scores relative to the mean and standard deviation of the unimpaired group. We reported adjusted mean differences between groups or regression coefficients (β), 95% confidence intervals and p-values. Statistical significance was set at α=0.05. Analyses were performed using R.

## Results

### Participants

The analytic sample included 122 community-dwelling Spanish-speaking Latino older adults, of whom 67% were female (n = 82). Participants had a mean age of 65 years (IQR: 60-69) and an average of 12 years of formal education (IQR: 8-16). The sample was predominantly cognitively unimpaired and without clinically significant depressive symptoms, with median MMSE and GDS scores of 27 (IQR: 25–29) and 5 (IQR: 2–8), respectively. However, 16 participants (13%) were classified as cognitively impaired, and 18 (14.7%) had GDS scores greater than 10, consistent with clinically significant depressive symptoms. Additional demographic and clinical characteristics of the sample are presented in Table 1.

**Table 1.** Sample demographics.

| Characteristic | N = 122<sup>1</sup> |
| --- | --- |
| Age (years) | 65 (60, 69) |
| Female sex (N, %) | 82 (67%) |
| Education (years) | 12 (8, 16) |
| Cognitively impairment (N, %) | 16 (13%) |
| MMSE score | 27 (25, 29) |
| LAS-PACC5 score | 0.09 (-0.62, 0.62) |
| GDS score | 5 (2, 8) |
| Plasma Aβ42/40 ratio<sup>2</sup> | 1.72 (1.40, 2.01) |
| Plasma brain-derived p-tau217<sup>2</sup> | 11.07 (10.89, 11.46) |
| Plasma NfL<sup>2</sup> | 11.51 (11.19, 11.97) |
| Plasma GFAP<sup>2</sup> | 12.95 (12.60, 13.44) |

<sup>1</sup>Median (Q1, Q3); N (%)
<sup>2</sup>Log-transformed and normalized plasma levels

### Plasma biomarkers, cognition and depressive symptoms

Impaired participants exhibited higher levels of plasma brain-derived p-tau217 (β = -0.75 [-1.39, - 0.10], p = 0.023) and GFAP (β = -0.57 [-1.07, -0.07], p = 0.026) compared to unimpaired participants (Figure 1). However, no differences between cognitive groups were observed for Aβ42/40 (β = -0.03 [-0.32, 0.38], p = 0.876) or NfL (β = -0.39 [-0.81, 0.04], p = 0.076).

**Figure 1.**
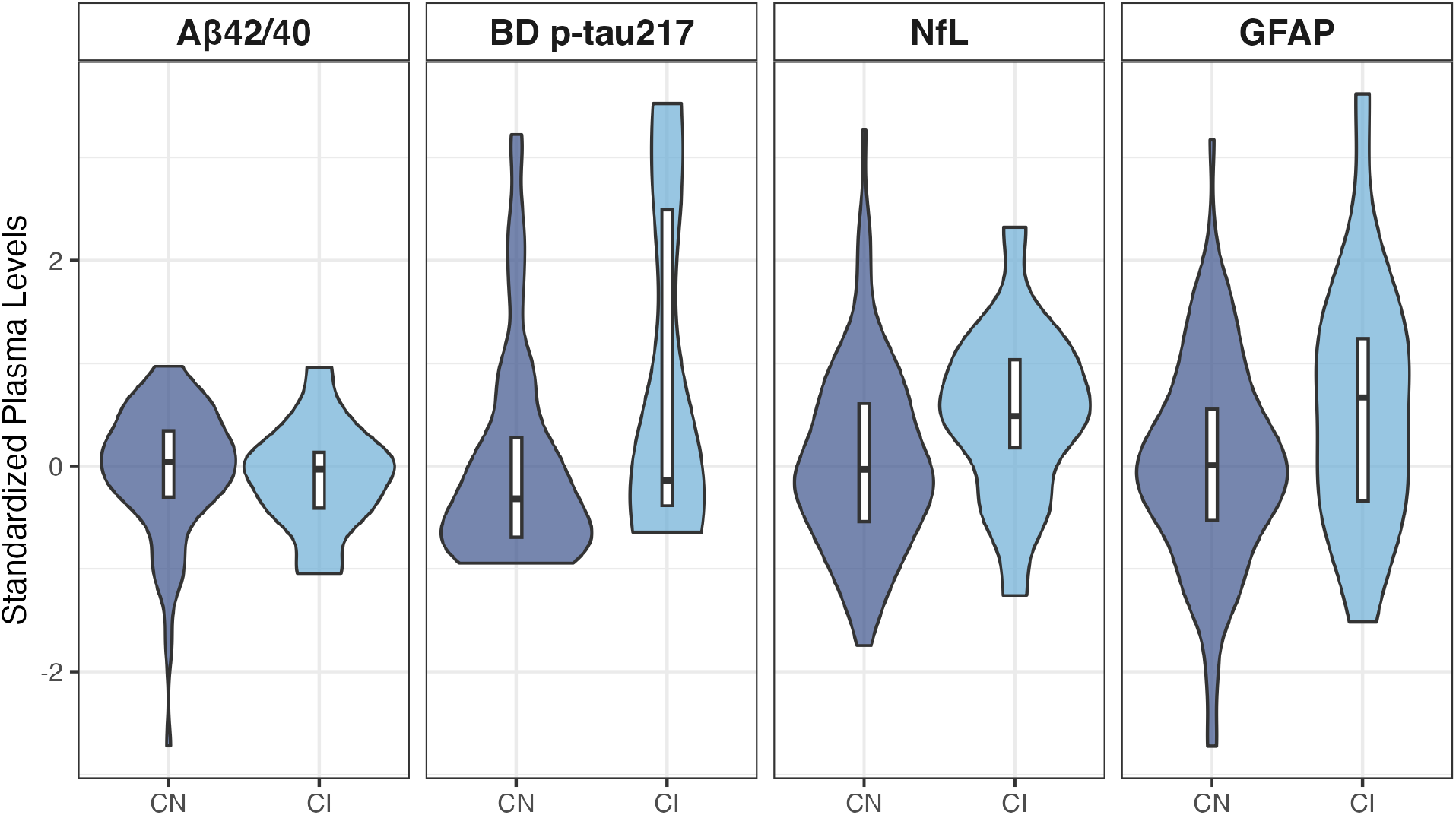
Differences in plasma biomarkers between cognitive groups. Violin plots showing the distribution of plasma biomarkers between cognitively normal (CN) and cognitively impaired (CI) participants. Box plots provide the median and IQR. Differences between groups were observed for plasma brain-derived (BD) p-tau217 (β = -0.75, p = 0.023) and GFAP (β = -0.57, p = 0.026), using analysis of covariance (ANCOVA) adjusted for age, sex and years of education.

Significant associations were observed between select plasma biomarkers and cognitive performance (Figure 2). Lower LAS-PACC5 scores, reflecting poorer global cognitive performance, were associated with higher plasma levels of brain-derived p-tau217 (β = −0.21 [-0.34, -0.08], p = 0.001) and GFAP (β = −0.24 [-0.40, -0.07], p = 0.006). In contrast, plasma Aβ42/40 (β = 0.06 [-0.19, 0.31], p = 0.633) or NfL (β = −0.11 [-0.31, 0.09], p = 0.287) were not significantly associated with LAS-PACC5 performance. A different pattern emerged for depressive symptoms. Higher GDS scores, indicating greater depressive symptom burden, were significantly associated with higher plasma NfL levels (β = 1.29 [0.01, 2.57], p = 0.048), whereas no significant associations were observed between GDS scores and Aβ42/40 (β = -0.52 [-2.12, 1.08], p = 0.522), brain-derived p-tau217 (β = −0.08 [-0.94, 0.77], p = 0.851) or GFAP (β = −0.17 [-1.27, 0.94], p = 0.763). Together, these findings demonstrate distinct cross-sectional associations across the biomarkers examined, with p-tau217 and GFAP associated with cognitive performance and NfL associated with depressive symptoms.

**Figure 2.**
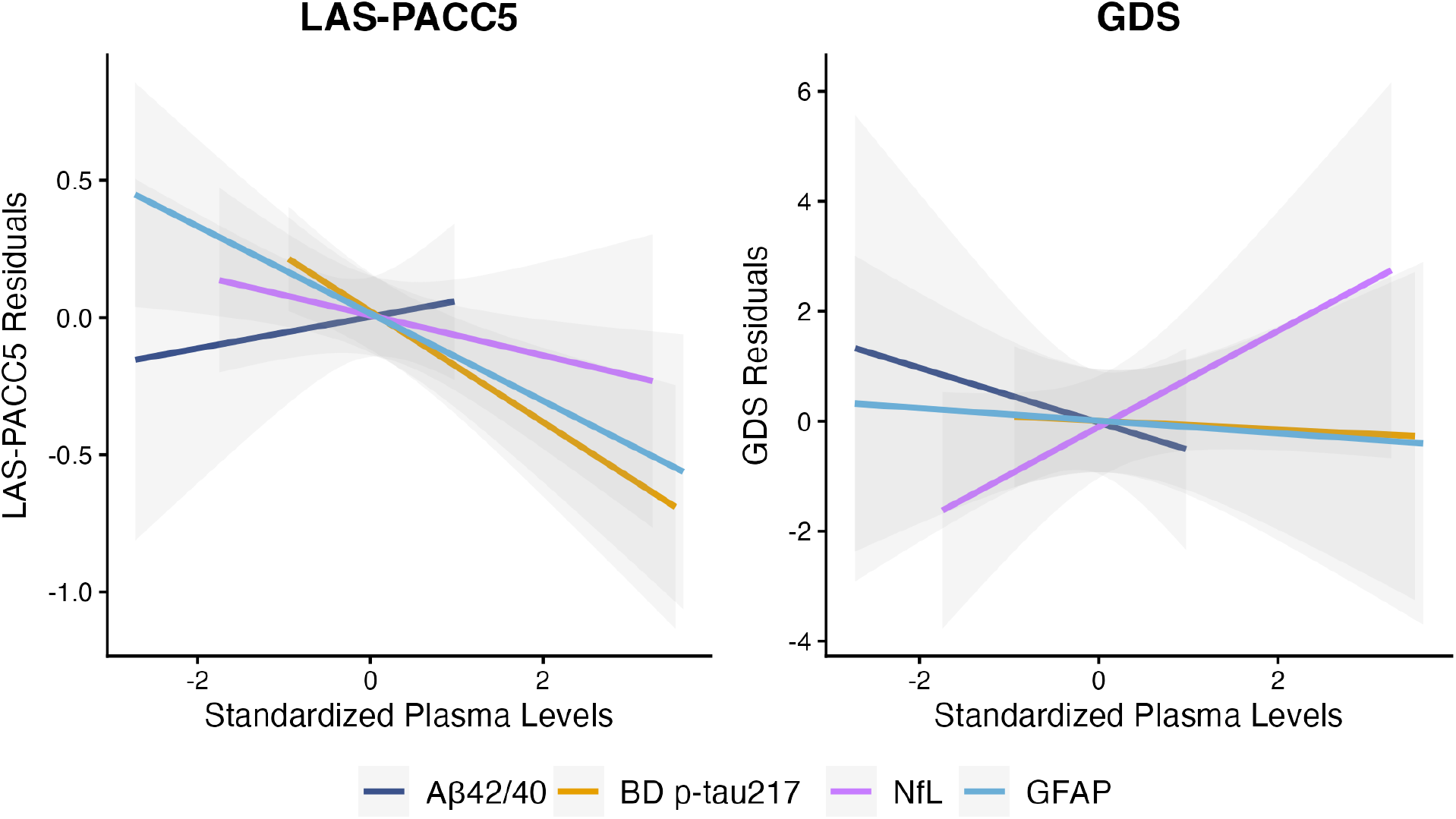
Associations between plasma biomarkers and LAS-PACC5 and GDS scores. Regression plots showing the associations of each plasma biomarker (colored) with LAS-PACC5 and GDS. The y-axis represents residuals from models adjusted for age, sex and years of education. Lower LAS-PACC5 scores were associated with greater plasma brain-derived p-tau217 (β = −0.21, p = 0.001) and GFAP (β = −0.24, p = 0.006) but not with Aβ42/40 or NfL. In turn, higher GDS scores were associated with greater plasma NfL (β = 1.29 [0.01, 2.57], p = 0.048) but not with Aβ42/40, brain-derived p-tau217 or GFAP.

## Discussion

In this community-based sample of Spanish-speaking Latino older adults, we found distinct associations between plasma biomarkers and cognitive and emotional functioning. Higher brain-derived p-tau217 and GFAP were associated with poorer global cognitive performance on the LAS-PACC5, while Aβ42/40 or NfL were not significantly associated with cognition. These findings add to growing evidence supporting blood-based biomarkers as minimally invasive indicators of biological processes associated with cognitive aging and AD[15,16]. The association between higher p-tau217 and poorer cognition is notable given evidence linking plasma p-tau217 with AD-related amyloid and tau pathology[16], while the association with GFAP may reflect astrocytic activation and related processes. Importantly, these associations were observed in a relatively young, community-dwelling sample with generally preserved cognitive functioning, suggesting that blood-based biomarkers may capture meaningful biological variability among Spanish-speaking Latino older adults without substantial global cognitive impairment.

A different pattern of associations emerged for depressive symptoms. Greater depressive symptom burden was associated with higher plasma NfL, whereas Aβ42/40, brain-derived p-tau217, and GFAP were not significantly associated with depressive symptoms. Because NfL is a marker of axonal injury that is elevated across a range of neurological and neurodegenerative conditions, this association may reflect broader processes related to brain health rather than AD-specific pathology [17]. This finding is particularly relevant given the growing recognition of behavioral and emotional changes as important features of cognitive aging and potential indicators of dementia risk[18,19]. These factors may be especially important in populations disproportionately exposed to socioeconomic adversity and other social determinants of health, which may contribute to both depressive symptoms and increased risk of cognitive decline and dementia. Together, these findings underscore the importance of considering emotional and social factors alongside biological markers when examining pathways to cognitive aging and dementia. The absence of an association between NfL and LAS-PACC5 performance, together with associations of p-tau217 and GFAP with cognition, suggests that these biomarkers may capture partially distinct biological processes relevant to cognitive and emotional functioning in Spanish-speaking Latino older adults.

Several limitations should be considered. First, the relatively small sample size limited statistical power and the precision of effect estimates, while also restricting our ability to examine potential heterogeneity across age, sex, education, country or region of origin, and other sociocultural and health characteristics. Second, the cross-sectional design precludes conclusions about the temporal or causal relationships between biomarker levels, cognitive functioning, and depressive symptoms. Longitudinal studies are needed to determine whether biomarker levels are associated with subsequent changes in cognition or depressive symptoms. Third, the relatively low burden of depressive symptoms in our sample may limit the generalizability of our findings to populations with more severe depressive symptomatology. Finally, because analyses were restricted to Spanish-speaking participants, the findings may not generalize to the broader, linguistically diverse Latino population represented in BLAST.

Overall, these initial findings support continued investigation of blood-based biomarkers for characterizing AD-related and neurodegenerative processes among Spanish-speaking Latino older adults. Larger samples and longitudinal follow-up are needed to determine whether baseline biomarkers predict subsequent cognitive or emotional changes. As BLAST expands, inclusion of Portuguese-speaking participants will broaden the cohort’s linguistic and cultural representation and allow examination of whether these biomarker associations are consistent across Spanish- and Portuguese-speaking Latino older adults.

## Data Availability

Anonymized clinical data are available upon request, subject to an internal review by Y.T.Q. to ensure that the participants confidentiality is protected. Data requests will be considered based on a proposal review and completion of a data sharing agreement, in accordance with the Mass General Brigham institutional guidelines. Please submit data requests to Y.T.Q.

## ACKNOWLEDGMENTS

The authors sincerely thank the BLAST study participants for their generous contributions of time and effort, which made this research possible. We also recognize the valuable support of former research staff members Nikole Bonillas-Felix, Lusiana Martinez, Randy Medrano, Nadeshka J. Ramirez-Perez, Isabel Solis, and Jorge Alcina in collecting data for the study.

## REFERENCES

1. Safiri, S. et al. (2024) Alzheimer’s disease: a comprehensive review of epidemiology, risk factors, symptoms diagnosis, management, caregiving, advanced treatments and associated challenges. Front Med (Lausanne) 11, 1474043. 10.3389/fmed.2024.1474043

2. Pahlke, S. et al. (2025) Blood-based biomarkers for detecting Alzheimer’s disease pathology in cognitively impaired individuals within specialized care settings: A systematic review and meta-analysis. Alzheimers Dement 21, e70828. 10.1002/alz.70828

3. Xiong, C. et al. (2024) Baseline levels and longitudinal changes in plasma Abeta42/40 among Black and white individuals. Nat Commun 15, 5539. 10.1038/s41467-024-49859-w

4. Hansson, O. (2021) Biomarkers for neurodegenerative diseases. Nat Med 27, 954–963. 10.1038/s41591-021-01382-x

5. Caviedes, A. et al. (2026) Blood-based AT(N) biomarkers for Alzheimer’s disease and frontotemporal lobar degeneration in Latin America. Nat Aging 6, 430–444. 10.1038/s43587-025-01061-3

6. Martinez, J.E. et al. (2025) Discussing the Interplay of Social Determinants of Health, Aging, and Alzheimer’s Biomarkers in U.S. Latinos. J Gerontol A Biol Sci Med Sci 80. 10.1093/gerona/glaf020

7. Association, A. (2026) Alzheimer’s disease facts and figures.. Alzheimers Dement 22, e71345

8. Rodriguez, M. et al. (2025) Associations between cognitive and clinical functioning and AD blood based biomarkers among Hispanic and WNH older adults. Alzheimers Dement. 25, e101390. 10.1002/alz70856_101390

9. Marquez, F. et al. (2025) Alzheimer’s disease plasma biomarkers are associated with cognitive performance among Hispanic/Latino adults. Commun Med (Lond) 6, 33. 10.1038/s43856-025-01295-7

10. Gonzales, M.M. et al. (2022) Blood biomarkers for cognitive decline and clinical progression in a Mexican American cohort. Alzheimers Dement (Amst) 14, e12298. 10.1002/dad2.12298

11. Jimenez, D.E. et al. (2020) Older Latino Mental Health: A Complicated Picture. Innov Aging 4, igaa033. 10.1093/geroni/igaa033

12. Gong, K. et al. (2025) High-sensitivity plasma proteomics reveals disease-specific signatures and predictive biomarkers of Alzheimer’s disease phenotypes in a large mixed-dementia cohort. Mol Neurodegener 20, 120. 10.1186/s13024-025-00909-x

13. Ramirez-Perez, N. et al. (2026) The Latin American Spanish PACC5 (LAS-PACC5): Advancing Cognitive Assessment in Latinos. Alzheimers Dement (Amst) 18, e70311. 10.1002/dad2.70311

14. Yesavage, J.A. (1988) Geriatric Depression Scale. Psychopharmacol Bull 24, 709–711

15. Hampel, H. et al. (2023) Blood-based biomarkers for Alzheimer’s disease: Current state and future use in a transformed global healthcare landscape. Neuron 111, 2781–2799. 10.1016/j.neuron.2023.05.017

16. Buckley, R.F. et al. (2026) Prognostic Value of Blood-Based P-Tau217 Levels for Progression to Cognitive Impairment. JAMA 336, 950–959. 10.1001/jama.2026.12556

17. Bellelli, F. et al. (2025) The relationship between neurofilament light chain and depressive symptoms according to cognitive status. Alzheimers Dement (N Y) 11, e70048. 10.1002/trc2.70048

18. Alcina, J. et al. (2025) Mild behavioral impairment and its relationship to cognition in community-dwelling older adults from the Boston Latino Aging Study. Alzheimers Dement (Amst) 17, e70121. 10.1002/dad2.70121

19. Elefante, C. et al. (2026) Mild Behavioral Impairment and Plasma Biomarkers of Neurodegeneration as Predictors of Cognitive Decline in Geriatric Patients With Psychiatric Disorders. J Geriatr Psychiatry Neurol 39, 413–425. 10.1177/08919887251394747

